# The effects of stress on the driving abilities of paramedic students: a pilot, simulator-based study

**DOI:** 10.1101/19003491

**Authors:** Trevor Hines Duncliffe, Brittany D’Angelo, Michael Brock, Cal Fraser, Jake Lamarra, Nick Austin, Matt Pusateri, Alan M. Batt

## Abstract

**Background:** Previous research has suggested that stress may have a negative effect on the clinical performance of paramedics. In addition, stress has been demonstrated to have a negative impact the driving abilities of the general population, increasing the number of driving errors. However, to date no studies have explored stress and its potential impact on non-clinical performance of paramedics, particularly their driving abilities.

**Methods:** Paramedic students underwent emergency driving assessment in a driving simulator before and after exposure to a stressful medical scenario. Number and type of errors were documented before and after by both driving simulator software and observation by two observers from the research team. The NASA Task Load Index (TLX) was utilised to record self-reported stress levels.

**Results:** 36 students participated in the study. Following exposure to a stressful medical scenario, paramedic students demonstrated no increase in overall error rate, but demonstrated an increase in three critical driving errors, namely failure to wear a seatbelt (3 baseline v 10 post stress), failing to stop for red lights or stop signs (7 v 35), and losing control of the vehicle (2 v 11). Self-reported stress levels also increased after the clinical scenario, particularly in the area of mental (cognitive) demand.

**Conclusion:** Paramedics are routinely exposed to acute stress in their everyday work, and this stress could affect their non-clinical performance. The critical errors committed by participants in this study closely matched those considered to be contributory factors in many ambulance collisions. These results stimulate the need for further research into the effects of stress on non-clinical performance in general, and highlight the potential need to consider additional driver training and stress management education in order to mitigate the frequency and severity of driving errors.

**Key points:**

- Paramedics are exposed to stressful clinical scenarios during the course of their work
- Many critical and serious clinical calls require transport to hospital
- Ambulance crashes occur regularly and pose a significant risk to the safety and wellbeing of both patients and paramedics
- This simulated clinical scenario followed by a simulated driving scenario has highlighted that stress appears to affect driving abilities in paramedic students
- The findings of this study, although conducted in paramedic students in simulated environments, highlight the need to further investigate the effects of stress on driving abilities among paramedics

## Introduction

Paramedic practice is enacted in inherently stressful environments and contexts. Sources of such stress include high acuity patients that decompensate quickly, performance of high-intensity procedures, a lack of personal time, and a disruption of social support (LeBlanc, 2009). Paramedics are regularly subjected to emotionally challenging work (Williams), violence (Bigham et al., 2014; Boyle and Wallis, 2016) occupational injury (Maguire et al., 2014), poor physical health status (MacQuarrie et al., 2018; Sheridan, 2019), and the effects of shift-work (Patterson et al., 2012). In addition, operational experiences such as attending hoax calls, missing meal breaks, and interacting with the public appear to influence psychological health, anxiety and stress in paramedics (Brough, 2004)

The literature on stress and performance is ambiguous and conflicting; some studies demonstrate no impact or even improvement of task performance, while other studies have demonstrated a negative impact on performance while stressed. For example, some research concludes that stress responses are associated with impairments in memory, attention, and decision-making abilities (Eysenck et al., 1987; Lieberman et al., 2002) while there is conflicting evidence that illustrates decision-making and memory in emergency workers are unaffected by stress (LeBlanc and Bandiera, 2007; Regehr et al., 2008). A number of previous studies demonstrated decreased performance on complex cognitive tasks, deterioration in communication skills, increased errors in medication calculations, and an increase in documentation errors among paramedics when stressed (LeBlanc et al., 2012, 2005). These results suggest that cognition and memory in paramedics could be affected by acute stress.

Research also illustrates that more experienced paramedics (the population in most of these studies) are not unaffected by stress but likely have developed more effective coping strategies to deal with stress (LeBlanc et al., 2012, 2005). This suggests that less experienced paramedics, including paramedic students and recent graduates, may be more susceptible to impaired performance when stressed than their more experienced counterparts, as they have not yet developed these coping strategies. In light of the potential relationship between stress and *clinical* performance, it is surprising that relatively little remains known about the effects that stress has on the *non-clinical* performance of paramedics. If stress does in fact impair other aspects of non-clinical performance, it could result in significant consequences for patient care, patient safety, and provider safety.

One of the core non-clinical aspects of the paramedic role is the driving component of traditional emergency ambulance work. Studies in the general public have demonstrated an association between stress and impaired driving, resulting in collisions (Rowden et al., 2011). Factors including adverse road conditions, other drivers, personality type, time constraints, and occupational stress have previously been linked to an increase in critical driving errors in the general public (Cartwright et al., 1996). Stressors in other domains of life such as work stress, daily hassles, and mental health issues can also adversely affect driving errors, concentration, and traffic violations (Rowden et al., 2011). Therefore, it is not unreasonable to assume that similar stressors may affect experienced drivers such as paramedics, and indeed it would be presumptuous to declare that paramedic driving ability is not affected by stress in the absence of any evidence. However, there is some evidence to suggest that paramedics possess higher awareness of driving threats when compared to members of the public, which may mitigate some of these effects (Pammer et al., 2018). Despite this suggestion, there remains an absence of research into the effects that stress may have on the driving performance of paramedics. For example, the majority of ambulance collisions occur while driving under emergency driving conditions with lights and sirens on during emergency calls (Murray and Kue, 2017; Watanabe et al., 2019) - a source of stress response in paramedic practice. It however remains unclear whether such collisions are the fault of the driver of the ambulance, or drivers of other vehicles (Kahn et al., 2001; Saunders, 1994). Given our lack of understanding of these collisions, and their potential implications for patient and paramedic safety we posit the potential causes of these collisions warrant further investigation.

However we face a challenge when we attempt to report statistics related to ambulance collisions. The data are elusive, and are highly dependent on the definition of “collision” used by each agency (which itself often remains unclear or unreported). Although this may seem trivial, it prevents researchers, policy-makers and end-users from adequately understanding the importance of the issue. The USA appears to have addressed some of these barriers through the development of a national reporting database. This database reported an annual average of 4,500 collisions that involved an ambulance, which resulted in an average of 33 fatalities and 2,600 injuries annually. Almost 60% of these collisions occurred during the course of emergency driving (National Highway Traffic Safety Administration, 2011a, 2011b; Smith, 2015). Figures for Canada remain largely unknown, with Alberta emergency vehicles involved in approximately 300 collisions per annum (Yasmin et al., 2014), and B.C vehicles involved in approximately 400 per annum (Tyakoff et al., 2014). UK figures present a similar challenge to locate; however, a search of grey literature indicated that the Welsh Ambulance Service experienced 735 emergency vehicle crashes between 2011 and 2016, and 150 occurred during emergency calls (BBC, 2011). Between 2009 and 2014, 148 ambulance collisions occurred in Berkshire, 310 in Hampshire, and 240 across Oxfordshire and Buckinghamshire (BBC, 2014) while 81 ambulances in the West Midlands crashed between 2012 and 2014 (Lumb, 2015). Even in light of the difficulty obtaining such data, the volume of events that occur across these jurisdictions suggest there is an onus on the community to investigate potential contributory factors involved in these collisions, and take steps to mitigate their impact.

It is important to clarify that we do not seek to claim stress as the sole, or even the most important, contributory factor that may affect the non-clinical performance of paramedics. A myriad of factors including fatigue, health status, and burnout may all influence paramedic performance (MacQuarrie et al., 2018; Ramey et al., 2019; Sofianopoulos et al., 2011). In fact, research demonstrates that acute stress is related to other health symptoms, such as fatigue, burnout, and post-traumatic symptoms (van der Ploeg, 2003) which further supports the need to investigate the potential impact of acute stress on performance. Therefore, the role of acute stress during the emergency driving phase of paramedic practice, before and after a stressful call, is the focus of our study. We aimed to examine the effects of acute stress on the number of critical driving errors made by paramedic students, and we hypothesized that they would demonstrate an increase in critical driving errors after the completion of a stress inducing clinical scenario.

## Methods and materials

### Participants

Participants were recruited from a convenience sample of first and second year paramedic students at [redacted for peer review], Ontario, Canada. All participants had a minimum of a valid G class licence (car licence) and knowledge of the Ontario Traffic Act and Ambulance Act. Participants were excluded from this study if they did not meet the above criteria.

### Ethics and consent

Ethical approval was received from [redacted for peer review] Research Ethics Board (protocol S 17-10-02-1). Participation was voluntary and all data were anonymized immediately following the simulation to maintain confidentiality. Participants were informed that they could withdraw their consent at any stage of the process prior to anonymization.

### Setting

This study was conducted in the province of Ontario, Canada. Paramedics in Ontario are generally staffed two per ambulance or solo response on a response vehicle. When they are double-crewed, they share both driving and patient care duties. The Highway Traffic Act (HTA) in Ontario governs the operation of motor vehicles on public roads, and provides for four primary exemptions (although there are more) for drivers of emergency vehicles:

1. Emergency vehicle drivers can proceed through a red light, after making a full stop and ensuring all traffic has stopped for them.
2. Emergency vehicle drivers are allowed to pass a streetcar on the left, because streetcars cannot pull over to the right.
3. Fire trucks are permitted to drive in the far left lane of the Don Valley Parkway, Gardiner Expressway and the 400 Series Highways
4. Emergency vehicle drivers can exceed the speed limit, as long as the lights and siren are activated (for ambulances: while responding to an emergency call or being used to transport a patient or injured person in an emergency situation).

(Ontario Ministry of Transport, 1990)

In order for these exemptions to be utilised, the vehicle must have flashing red/white/blue lights and siren activated simultaneously, and it must be a true emergency. Emergency vehicle drivers must operate the emergency vehicle with due regard for the safety of others, on and off the road (Krkachovski, 2008). In addition, emergency vehicles are granted certain additional privileges under the HTA, such as the ability to drive on closed highways, and use of communication devices. They are not permitted to drive the wrong direction on streets, and they are not exempted from coming to a complete stop at stop signs before proceeding through the intersection. Paramedic students receive education related to these privileges and exemptions.

### Materials – driving simulation

The simulated driving scenario was conducted using a PatrolSim ambulance driving simulator [*Version 4, L3 Training Solutions, Salt Lake City, UT*] based at [redacted for peer review]. This is an open-cabin simulator which simulates driving conditions in both emergency and non-emergency modes, visually and aurally (see Figure 1). It does not have the ability to simulate motion. While the evidence related to validity of driving simulators is mixed (Mullen et al., 2011), the use of a driving simulator in this pilot study was designed to minimize ethical concerns and risks related to the operation of real vehicles by participants, particularly in the post-stress phase. While driving simulators are a poor choice for the assessment of real world driving (for example as a substitute to road testing), they are generally considered a valid tool for researching driving (Mullen et al., 2011). The simulator provides navigation directions on screen using large arrows which denote route of travel similar to that of commonly used global positioning system car navigation units. This simulator is also capable of recording a number of critical driving errors. Participants were initially provided with an orientation and test run in the simulator prior to any data collection (driving or clinical scenario) in order to provide them with the opportunity to understand the simulator functions. Participants were then given a brief overview of the HTA and Ambulance Act to ensure that all participants understood the rules of the road. A baseline driving assessment was then performed whereby each participant drove the simulated ambulance under emergency driving conditions from a known starting position to a denoted endpoint. Pre-programmed, standardised “*EMS_City_2”* and “*EMS_City_3”* routes were used, each consisting of a 2-3 minute drive with a set start and end location. These routes are artificial simulated driving routes, and do not represent or mimic a ‘real world’ driving route that students would be familiar with. The “baseline” drive and “post-stress” drive were standardized for each participant.

**Figure 1.**
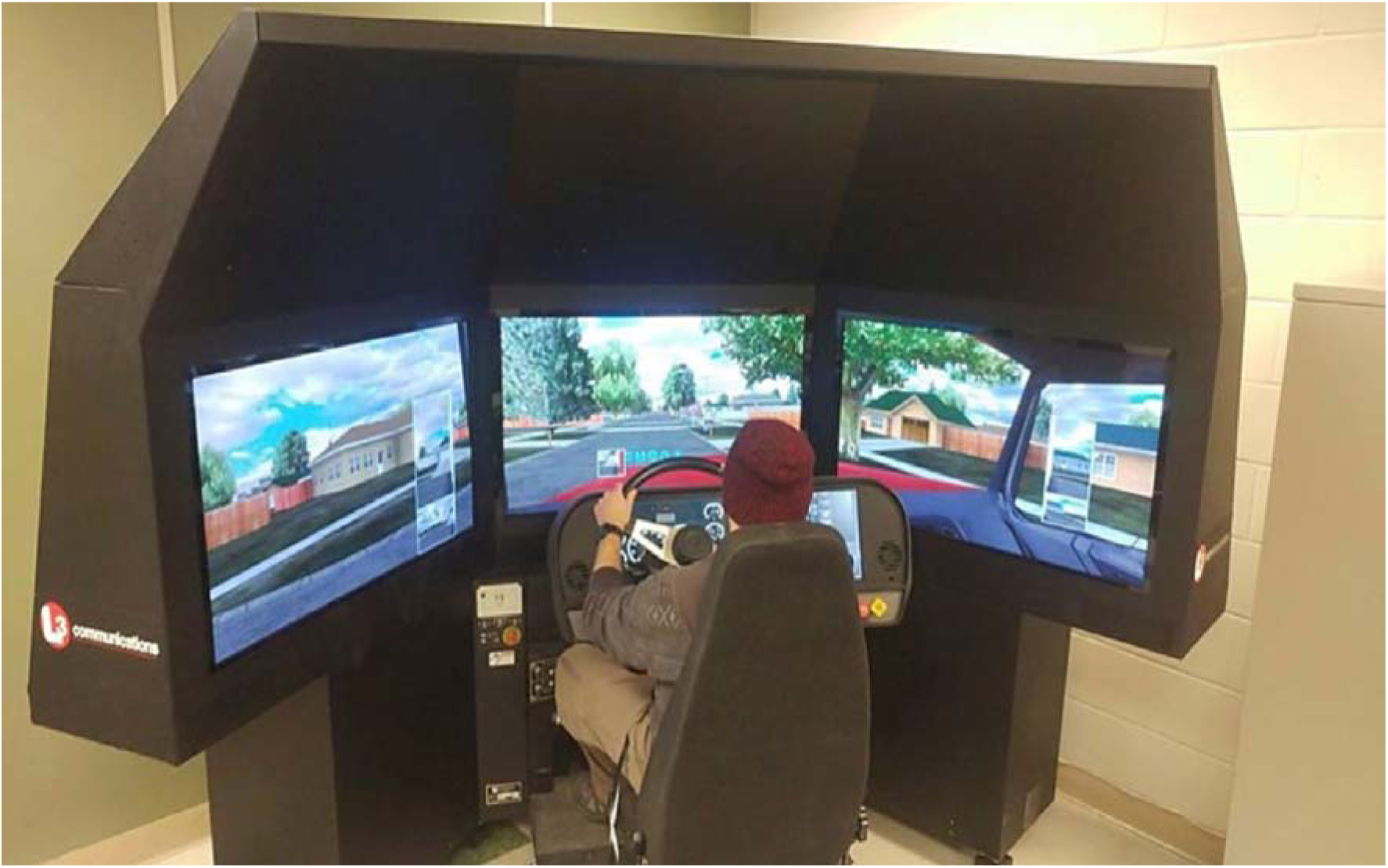
PatrolSim ambulance driving simulator [Version 4, L3 Training Solutions, Salt Lake City, UT].

Data were collected using two complementary approaches. First, a computer generated log of driving errors was captured by the PatrolSim software. These errors included: failure to wear a seatbelt (not exempted under HTA); exceeding the posted speed limit (exempted with conditions under the HTA); and collisions. Second, critical errors not recognized by the driving simulator were recorded by the researchers, and included: failure to stop at a stop sign (controlled intersection; NOT exempted under HTA), running a red light (controlled intersection; exempted with conditions under HTA), and loss of vehicle control. We defined loss of control of the vehicle as *“the act of the driver being unable to successfully maneuver the vehicle, resulting in the simulated vehicle skidding off the road surface and requiring the vehicle to come to a stop to regain control”*.

### Materials – stress inventory

Once the participants had completed each drive, they completed a self-reported stress-inventory test on a supplied tablet device. The National Aeronautics and Space Administration Task Load Index (NASA-TLX) is a widely used instrument to assess overall subjective workload, and has previously demonstrated accurate test-retest reliability, validity and sensitivity (Hoonakker et al., 2011), as well as being easy to administer and analyse via an official NASA developed mobile app. The NASA-TLX is a multidimensional instrument comprised of six subscales designed to measure workload; mental demand, physical demand, temporal demand, performance, effort, and frustration (Hart and Staveland, 1988). The NASA-TLX requires participants to weigh and rate these categories on a sliding scale, on a 20-point scale ranging from “Low” to “High”, resulting in a report of the overall workload burden experienced by the participant on a scale from 0 to 100. The participants repeated the NASA-TLX after each drive in the simulator to compare their perceived workload burden between drives.

### Materials - clinical simulation

Participants individually completed a 12-minute, stressful clinical simulation which involved the resuscitation of a paediatric patient. Paramedics have previously reported increased stress and anxiety when caring for paediatric patients, and they themselves perceive this to be an important contributor to patient safety events (Guise et al., 2017). This scenario was reviewed by faculty members to ensure clinical accuracy and to predict its ability to induce stress in paramedic students. All participants completed the same scenario. Immediately following the scenario, the individual was required to drive the simulator under emergency driving conditions from another standardised start to end point. They were also required to complete the NASA-TLX for a second time. Participants were then given the opportunity to debrief after their participation to address any concerns, and were informed of additional support services.

### Data Analysis

“Baseline” and “post-stress” driving errors were compared using results from the driving simulator’s computer-generated report of critical errors as well as critical driving errors recorded by two observers. “Baseline” and “post-stress” perceived levels of stress were compared using results from the NASA-TLX. Descriptive statistics are used to illustrate data. We now report on critical driving errors that occurred under emergency driving conditions, namely failure to wear a seatbelt, failure to stop (red light or stop sign at a controlled intersection), and loss of control of vehicle.

## Results

A total of 36 participants completed the study with 29 first year students (81%) and 7 second year (19%). Critical driving errors occurred more often “post-stress” than at baseline. Failure to wear a seatbelt, failure to stop for red lights or stop signs, and loss of control of the vehicle all demonstrated an increase after exposure to stress compared to “baseline” (see Figure 1).

**Figure 1.**
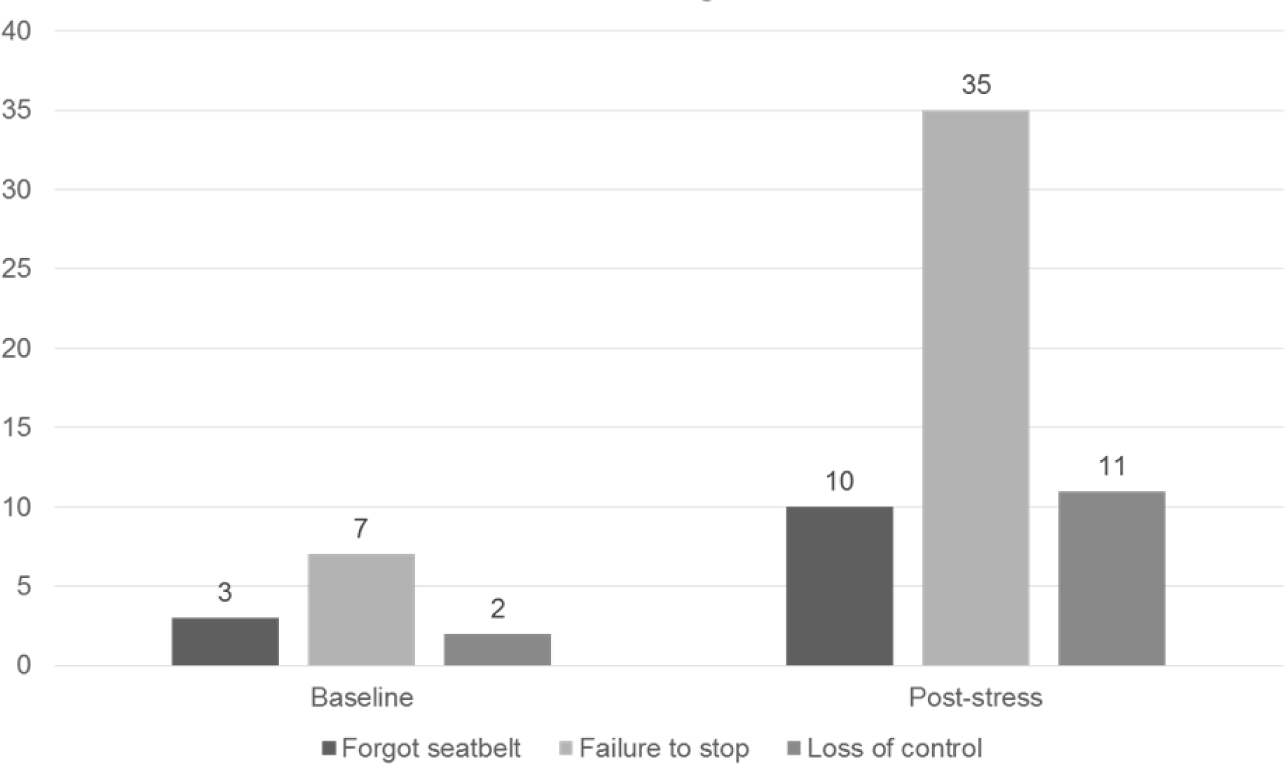
Critical driving errors at baseline and post-stress.

Participants reported an increased level of stress during the “post-stress” drive (overall weighted stress score of 67.59/100) compared to “baseline” drive (overall weighted stress score of 63.81/100) via the NASA-TLX (Figure 2). When stress scores were broken down into specific categories, it was found that on average participants rated “mental demand” the highest source of stress (73.1/100), followed by “frustration” (64.86/100), “effort” (63.47/100), “temporal (time) demand” (61.39/100), performance demand” (54.58/100), and finally “physical demand” (26.39/100). No participants reported experiencing simulator sickness.

**Figure 2.**
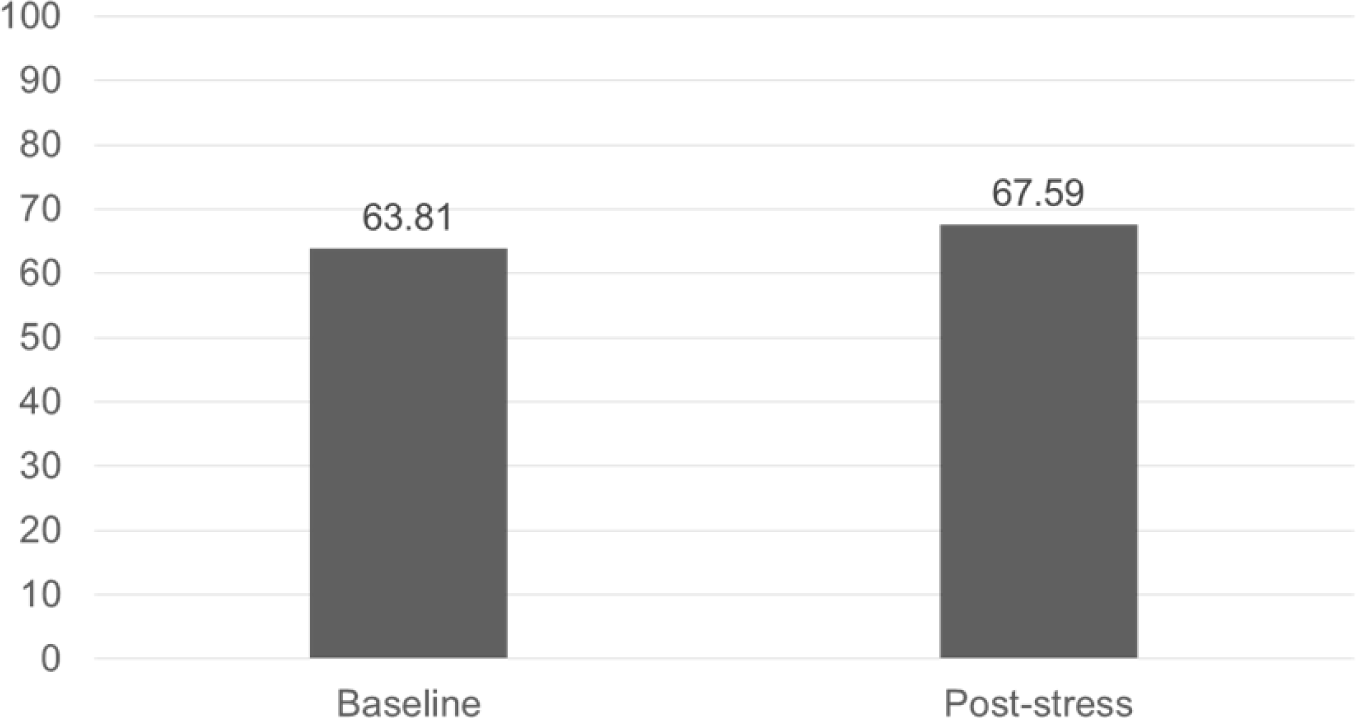
Mean NASA-TLX reported stress scores baseline and post-stress.

## Discussion

The results of this pilot study demonstrate that paramedic students make more critical driving errors when stressed, namely failure to wear a seat belt, failure to stop at stop signs or red lights (intersections), and loss of control of the vehicle. The types of errors made after stress exposure suggest that paramedic students may drive in a manner that is less defensive and more unsafe during transport after stressful calls. Participants also reported higher levels of stress after a stressful clinical scenario, in particular in the domain of mental (cognitive) demand. Our findings suggest that acute stress may affect the non-clinical performance of paramedic students. These findings are important for the community to reflect on (bearing in mind their limitations), given the potential consequences for both paramedics and patients.

Participants reported increased mental demand on the NASA-TLX during this study. A possible cause for the rise in the specific critical errors reported in this study may be due to cognitive overload from the stressful stimuli that were induced. We are all constantly overloaded with perceptual information in our daily lives and activities. Our ability to process this volume of information is limited, and under stressful conditions the cognitive system can become overloaded, decreasing resources devoted to attention, and therefore our ability to concentrate and perform a specific task (LeBlanc, 2009). This increased mental demand may potentially contribute to lack of awareness or risky decision making leading to critical errors when operating an ambulance under emergency driving conditions.

The particular critical errors demonstrated by participants in this study were interesting to note in that they appear related to driving while stressed under emergency driving conditions. This is an interesting observation, as fatal ambulance collisions are more prevalent when the lights and sirens are activated during emergency driving (Kahn et al., 2001). For example, we discovered that participants were 3.3 times less likely to wear their seatbelt when driving “post-stress”. Failure to wear a seatbelt while driving is inherently dangerous, and results in significantly higher mortality rates when compared to collisions where seatbelts were worn (Abbas et al., 2011). Unfortunately multiple examples can be identified via media reports of paramedics and patients who have been seriously injured or killed due to lack of restraint in a collision. Participants also demonstrated an increase (post-stress) in failure to stop at stop signs or red lights (controlled intersections) before exercising exemption (applicable to red lights) and proceeding through them under emergency driving conditions. Failure to stop is one of the leading causes of ambulance collisions in the USA (Retting et al., 2003). Injuries and fatalities are more likely in collisions involving missed stop signs or red lights than in other crash types (Retting et al., 2003; Yasmin et al., 2014). In addition, most fatal ambulance crashes in the USA occur while going through an intersection with the emergency lighting and sirens activated (Kahn et al., 2001). Our findings, combined with the evidence from the literature, suggests that activation of lights and sirens alone may be insufficient measures to prevent collisions at controlled intersections. Reinforcing existing advice that emergency vehicle operators come to a complete stop at stop signs and red lights under emergency driving conditions before proceeding appears appropriate in order to reduce these types of collisions (Krkachovski, 2008).

Given the occupational stress and time constraints regularly encountered by paramedics (LeBlanc et al., 2012), and in light of our findings, we suggest that the link between stress and non-clinical performance is worthy of further exploration. Specifically to driving performance, existing guidance related to driving under emergency conditions may need to be revisited or clarified. Even 20 years ago, Whiting et al. suggested that those with less experience and perhaps education could benefit from additional training in an attempt to reduce the number of ambulance collisions (Whiting et al., 1998). For example, the provision of advanced driver training may mitigate some of these forms of collisions through earlier recognition and anticipation of potential issues, leading to “expert drivers” (Pammer et al., 2018). Given the potential negative impact of stress on performance educators might also consider avenues for training learners in stress management (LeBlanc, 2009).

Finally, although not a direct outcome of our study, our preliminary work focused on gathering data and identifying literature to support and inform our study. Efforts to source these data presented challenges, which we have discussed elsewhere (redacted for peer-review). In particular, the lack of easily accessible statistics on the incidence of emergency vehicle and ambulance collisions presented a challenge. This is caused by a lack of national reporting databases, and makes it difficult for researchers and policymakers to base recommendations on appropriate evidence. It also results in an observable lack of standardization; the definition of ‘collision’ for example was often ambiguous, undefined or conflicted with a definition used elsewhere. This lack of standardization poses a challenge when attempts are made to quantify the scale and impact of the issue. Future endeavours to implement national or regional standardized reporting measures should seek to resolve this obstacle.

### Limitations

Our small sample size means our results were prone to error in statistical analysis, and thus we elected to discuss our results in terms of descriptive statistics only. The results may not accurately reflect driving abilities of all paramedic students in the program, and this reduces the external validity of the results. However, the findings of this pilot study are designed to stimulate discussion and provide a foundation for future, more robust research on the issue. In addition, the majority of participants were first year paramedic students with limited ambulance operations experience. Although the driving simulator is able to recreate the visual and auditory aspects of driving quite accurately, it is unable to recreate the physical forces (g-forces) and sensations of acceleration, deceleration, and turning. Participants reported this made it difficult to judge their speed, how hard they were braking, and how aggressively they were steering in the simulator. Therefore, our results cannot be directly translated to the driving of real vehicles; however, the underlying suggestion that stress appears to affect driving (non-clinical) performance is worthy of further investigation, and to date has not been well investigated in paramedics. The conduct of this study with real emergency vehicles would be fraught with issues surrounding ethical conduct and participant safety. On the other hand, future studies in this area would benefit from studying a larger population of clinically active paramedics through ethically appropriate means, as well as paramedic students and recent graduates. These future studies may yield results that are more applicable to the population who are affected by the issues highlighted in our results. Future research could also consider objective physiological markers of stress in addition to subjective self-reported stress approaches.

## Conclusion

Paramedics are subjected to stressful situations on a regular basis in their careers, which may affect both their clinical and non-clinical performance. One of the main non-clinical roles in emergency paramedic work is the driving of ambulances under non-emergency and emergency driving conditions. Our results demonstrate that paramedic students reported higher levels of stress after a stressful simulated clinical scenario. In a simulated driving environment they demonstrated an increase in critical driving errors after a stressful simulated clinical scenario. These critical errors represent potential contributory factors in ambulance collisions, namely failure to stop, failure to wear a seatbelt, and loss of control of the vehicle. The findings of this study should stimulate increased dialogue on the effects of acute stress on paramedic performance, its potential implications for both paramedics and patients, and further research into the effects of stress on driving ability and other non-clinical aspects of the paramedic role.

## Data Availability

Data are available on request

## Acknowledgments

The authors wish to thank Ms. Lauran Livingston for her contributions to the conduct of the study. We also wish to thank Mr. Mark Dwyer and Mr. Jason Constable for their assistance with the conduct of the study.

